# Identifying the practice patterns of optometrists in providing falls prevention management: A mixed-methods systematic review protocol

**DOI:** 10.1101/2024.05.15.24307394

**Authors:** Si Ye Lee, Khyber Alam, Jason Charng, Hamed Niyazmand, Jingyi Chen, Anne-Marie Hill

## Abstract

**Objective:** The objective of this systematic review is to synthesise the best available evidence for optometrists’ practice patterns in providing falls prevention management.

**Introduction:** Falls remain the main cause of injury-related hospitalisation and mortality in Australia and worldwide, significantly affecting older adults. The increased risk of comorbidities, including visual impairment in this cohort is linked to a higher incidence of falls. Despite being primary eye care practitioners, community optometrists may not consistently integrate falls prevention strategies into their practice. Furthermore, the extent to which they adhere to evidence-based recommendations for falls management remains unclear.

**Inclusion criteria:** The review will include optometrists, in regions where optometry is a regulated profession, and report their understanding and practice patterns in delivering falls prevention management to older community-dwelling adults. Qualitative, quantitative, and mixed methods studies will be eligible for inclusion. It is envisioned that most studies will be qualitative. Studies published in English and those published from 1980 onwards will be eligible for inclusion since published evidence for falls prevention began to increase sharply around this time.

**Methods:** The review will follow the JBI guidelines for mixed methods systematic reviews and will be developed and reported in accordance with PRISMA-P guidelines. Databases that will be searched are Excerpta Medica Database (Embase), Scopus, Cumulative Index to Nursing and Allied Health Literature (CINAHL) Complete, and OVID MEDLINE. Grey literature will be searched through Conference Proceedings Citation Index (Web of Science), Google Scholar, and ProQuest Dissertations databases. Two reviewers will independently conduct all screening and critical appraisal. The reviewers will screen all articles’ titles and abstracts retrieved from the searches to determine potential eligibility. All full-text articles considered relevant will then be assessed for final eligibility for inclusion. The final included articles will be assessed for methodological rigour using the JBI SUMARI critical appraisal tools, subsequently, all relevant data will be extracted. Discrepancies at any stage of the procedures will be resolved through discussion and consensus with a third senior researcher. A convergent integrated approach to synthesising and integrating the quantitative and qualitative data will be followed.

Review registration CRD42024539668

## Introduction

Falls remain Australia’s leading cause of injury hospitalisation and death, with the Australian Institute for Health and Welfare (AIHW) identifying that falls represented 43% of injury hospitalisations and 42% of injury deaths in 2023.^(1)^ Throughout 2021-22, falls-related injuries accounted for 233,000 hospital admissions, and in 2020-21 accounted for 5,800 deaths.^(1)^ Falls are a significant problem amongst older adults (aged ≥ 65 years old) since this cohort of the population accounts for 60% of falls-related hospitalisations and 94% of falls-related deaths.^(1)^ The economic cost associated with hospitalisations due to falls among older adults in Australia has been estimated at $2.3 billion annually.^(2)^ Globally, older adults experience an average of 0.67 falls annually, with population-based studies indicating that 10% of older adults have two or more falls per year.^(3)^ Worldwide, falls contribute to over 38 million DALYs (disability-adjusted life years) lost annually, resulting in greater years living with disability compared to transport injury, drowning, burns and poisoning combined.^(4)^

Optometrists, as primary eye care practitioners have the skills and knowledge to diagnose and manage various eye conditions that lead to visual impairment. Evidence has shown a strong correlation between visual impairment and an increased risk of falls and mortality.^(5)^ The prevalence of visual impairment poses a significant challenge in older adult populations, with statistics indicating that more than one in four older adults are affected.^(6)^ Older adults are at a greater risk of eye disease^(6–8)^ including chronic ocular problems such as refractive errors,^(9)^ cataracts,^(7)^ glaucoma,^(8)^ and age-related macular degeneration (AMD).^(8)^ Patients diagnosed with glaucoma often exhibit constricted peripheral vision as the disease progresses, leading to problems in safely monitoring the environment and consequently, an increased risk of falls.^(10)^ Other ocular conditions such as cataracts, cause a decrease in the patient’s visual acuity and contrast sensitivity over time, with falls occurring in 31% of patients while waiting for cataract surgery.^(11)^ Patients with AMD can experience reduced central vision, which impairs their mobility and can result in a higher prevalence (74%) of falls compared to older adults without AMD.^(12)^ Additionally, older adults frequently require specialty lenses due to refractive errors such as presbyopia.^(13)^ When presbyopia is corrected with multifocal spectacles the unique design may create peripheral distortion. This distortion may result in blurred vision when viewing floor-level objects, potentially exacerbating the risk of falls.^(13)^

The Australian Health Department recommends that all older adults undergo a visual assessment yearly.^(14)^ When older adults present for a vision assessment, they may only have a mild visual impairment. However, older adults may have multiple other risk factors for falls, including medications that reduce their balance, arthritis, prior fractures, mobility problems, and reliance on walking aids.^(15)^ Recognising the high incidence of falls and injuries among older adults, recently published World Falls Guidelines (WFG) recommend initiating falls prevention screening and management for older adults starting from the age of 65 years.^(9,15)^ The WFG recommends that all health professionals routinely ask about falls in their interactions with older adults to identify the risk of falls and proceed to appropriate management depending on the clinical presentation.^(15)^ Recommended management is to offer general falls prevention advice to all older adults, even those who are screened as being at low risk of falls. Annually, healthcare practitioners should prompt older adults to disclose if they have experienced one or more falls in the past year or exhibit other risk factors for falls. When older adults experience one or more falls, they should be offered targeted exercise or a multifactorial assessment depending on the level of risk identified.^(15)^ The WFG also offers vision-specific guidelines aimed at enhancing the scope of optometrists in delivering comprehensive falls prevention management for older adults. These guidelines delineate several key concepts and practice points to assist optometrists in conducting thorough assessments and interventions. Specifically, optometrists are advised to inquire about any visual impairments, assess visual acuity, contrast sensitivity, and depth perception, and examine peripheral vision to detect conditions such as hemianopia or neglect.^(15)^ Furthermore, the guidelines recommend updating any outdated prescription spectacles or provisioning new ones to older adults who do not possess them, as this can benefit their visual perception to lower the risk of falls. Further recommendations include advising patients with diagnosed cataracts to undergo cataract surgery on the initial eye and subsequent eye.^(15)^ The WFG recommends that clinicians inquire about older adults’ priorities and attitudes to inform decision-making and advice for falls prevention. Moreover, clinicians are recommended to promote shared decision-making and evidence-based care provision that involves patients in the process to enhance engagement and ensure effective risk-reduction strategies.^(15,16)^

Optometrists, being primary eye care practitioners, have the professional capacity to screen and provide appropriate falls prevention advice for all older adults who present to their practice, as well as specific visual advice if the older person has a visual impairment. However, there has been limited exploration regarding the role and falls prevention practice patterns of optometrists when managing older patients, despite studies suggesting that there is a benefit to involving optometrists in falls prevention management.^(16–19)^ Other allied health professionals (e.g. physiotherapists, occupational therapists, pharmacists) have extensively investigated how to deliver falls prevention management, with multiple systematic reviews published within these disciplines, exploring the effects of medication reviews,^(20)^ home modifications,^(21)^ education physical training,^(22)^ and exercise to help improve falls management.^(22–24)^ However, there is limited evidence of how optometrists could implement recommendations for falls prevention management into practice. Existing Optometry Australia guidelines provided by the Queensland University of Technology and the University of Bradford suggested refractive alterations in those with moderate-high risk of falls, and other recommendations include having a targeted case history revolving around previous falls.^(25)^ However, the guidelines lack comprehensive recommendations on the consistent implementation of falls prevention management. Furthermore, the guidelines do not incorporate guidance about referral pathways or co-management with other health practitioners, such as physiotherapists or occupational therapists. A recent study suggested that many optometrists consider the current guidelines as a reference, and their implementation of the guideline varies, based on the perceived importance and feasibility of implementing the recommended strategies to their practice.^(16)^

Given increasing falls rates worldwide, the WFG has emphasised the need for all health professionals to provide falls prevention management. Hence, the involvement of optometrists as primary practitioners who see a large proportion of the older adult population will be important in community settings. The objective of this review is to synthesise the best available evidence for optometrists’ practice patterns in providing falls prevention management. Findings from this review will inform the development of education around falls prevention management for optometrists and how this gap can be addressed with future recommendations for optometrist clinical practice.

A preliminary search of MEDLINE, PROSPERO, and Cochrane Database of Systematic Reviews found two ongoing systematic reviews, related to optometry and falls. The first review investigates what interventions can reduce falls risk in older adults with visual impairment.^(26)^ The second is investigating the prevalence of falls globally among individuals aged 18 to 65 with low vision.^(27)^ This systematic review will differ from these reviews since it will identify and synthesise the evidence for optometrists’ practice patterns in falls prevention management for older community-dwelling adults.

## Review Questions

1. What are the current practice patterns for optometrists in providing falls prevention management for older adults?
2. Do optometrists assess older adults for their risk of falls?
3. Do optometrists provide falls prevention advice or referral for older community-dwelling adults who present to their practice?
4. Are optometrists aware and motivated to provide falls prevention management for older patients as part of their routine ocular assessments?

## Methods

A mixed-methods systematic review will be conducted in accordance with the JBI guidelines, employing a convergent integrated approach for synthesising and integrating data.^(28)^

## Inclusion criteria

Following the JBI convergent integrated approach for a mixed-methods systematic review,^(28)^ the inclusion and exclusion criteria will be defined by “population”, “phenomenon of interest” and “context”.

### Population

The review will consider studies that investigate practice patterns of registered optometrists working within a community-based setting. Studies will be included if they report findings where optometrists, directly assess, and manage the older adult patients who present to the practice. In this review, registered optometrists will be defined as qualified eye care practitioners registered with a national health practitioner regulation agency such as (Australian Health Practitioner Regulation Agency AHPRA, General Optical Council United Kingdom, State Boards of Optometry United States and equivalent in other countries where optometry is a regulated profession) and trained in primary eye and vision care.^(29)^ No exclusion criteria based on the study participants’ characteristics such as years of work experience will be applied.

### Phenomenon of Interest

All studies that explore the practice patterns of optometrists when consulting with older adult patients, specifically their practice in falls prevention management will be considered for inclusion. Practice patterns will also extend to the optometrist conducting any form of falls prevention screening, assessment, management or providing advice or guidance to their patients. For the purposes of the review, older aged adults are defined using the MeSH description of adults aged 65 years and older.

### Context

Studies conducted in community settings where older adults undergo vision assessments will be included. Studies set in environments such as residential aged care facilities and hospitals will be excluded, as older adults in these settings receive specialised care from teams of professional staff. Studies where doctors or nurses provide eye care assessments or advice will also be excluded as the study seeks to understand the practice patterns of optometrists as primary care practitioners in the community.

## Types of Studies

This review will consider quantitative, qualitative, and mixed methods studies. Quantitative studies will include experimental, descriptive correlation, and quasi-experimental studies. Qualitative studies will include phenomenological, ethnographic, grounded theory, historical, and action research studies. Mixed method studies will only be considered for inclusion if data from the quantitative or qualitative components can be extracted separately. Other reviews will be excluded, but if there are any relevant articles from their reference lists, these articles will be included in the screening process. Studies published in English will be included. Studies published in English and those published from 1980 onwards will be eligible for inclusion since published evidence for falls prevention began to increase sharply around this time.

## Search strategy

The search will seek to identify both published and unpublished studies. The search strategy follows the 3-step method described previously by JBI.^(30)^ First, a limited search of MEDLINE and CINAHL Complete will be conducted, followed by an analysis of the text words found in the titles and abstracts, as well as the index terms and Medical Subject Heading (MeSH) terms used to describe the articles. Second, these results will subsequently assist to inform the development of a search strategy that will be tailored for each information source. MeSH keywords and index terms will be used to develop a full search strategy with the guidance of an experienced librarian. Third, the reference list of all studies selected for critical appraisal will be hand-screened for additional studies. An example of a search strategy for OVID MEDLINE is detailed in Appendix 1.

Following the search, all citations found will be loaded into COVIDENCE (Veritas Health Innovation, Australia), and duplicates will be removed. COVIDENCE is an online collaboration software platform designed to streamline the process of producing systematic and other literature reviews. Subsequently, two independent reviewers will screen titles and abstracts to assess their eligibility against the inclusion criteria for the review. Any potentially relevant studies will then be retrieved in full.

The full text of selected citations will undergo further assessment against the predetermined inclusion criteria by two independent reviewers. Any studies failing to meet these criteria will be excluded, with the reasons for exclusion documented and reported. Authors will be contacted to request additional information if clarification is needed. Should discrepancies between the reviewers arise, they will be resolved through discussion or with the involvement of a third senior reviewer. The search results and final study inclusion will be reported fully in the final review and depicted in a Preferred Reporting Items for Systematic Review and Meta-Analyses (PRISMA-20) flow diagram, ensuring transparency and clarity in the selection process.^(31)^

## Information Sources

The databases to be searched are the Excerpta Medica Database (Embase), Scopus, Cumulative Index to Nursing and Allied Health Literature (CINAHL) Complete, and OVID MEDLINE, and the search strategy will be adapted between the databases.

The search for unpublished studies and grey literature will be completed using the Conference Proceedings Citation Index (Web of Science), Google Scholar, and ProQuest Dissertations.

## Assessment of Methodological Quality

Quantitative studies (including quantitative components of mixed methods studies) will be assessed for methodological quality using the standardised critical appraisal tool set out by JBI SUMARI (found in appendix 4.1 on the JBI website).^(32)^ Qualitative studies, encompassing any qualitative components in mixed-methods studies, will be assessed for methodological quality utilising the standardised critical appraisal tool set out by JBI SUMARI (found in appendix 3.1 on the JBI website).^(33)^

The results of critical appraisal will be reported in narrative form and table format. All studies, regardless of their methodological quality assessment results, will undergo data extraction and synthesis. The methodological quality of the studies (low vs. high) will be considered at the time of data analysis.

## Data Extraction

Quantitative and qualitative data will be extracted from the included studies by two independent reviewers using COVIDENCE (Veritas Health Innovation, Melbourne, Australia) and the standardised JBI data extraction tool set out in JBI SUMARI (tool provided in appendix 8.1).^(28)^ The data extracted will include specific details about the study information such as title, authors, country of origin, language, study year, year of publication and journal, and study characteristics about the population, study methods, phenomena of interest, context and relevant outcomes about the research questions will also be extracted.

Quantitative studies will consist of descriptive, or analytic studies that provide information about magnitude and statistical significance. For descriptive studies, data extraction will comprise an average or a percentage that can profile the sample within the study. At the same time, analytic studies will examine the relationship between the variables where extracted data will involve all relationships relevant to the review question.^(28)^ Data that report the prevalence of falls prevention management, levels of knowledge about falls prevention management, attitudes and motivation on undertaking falls prevention management, prevalence of falls risk screening, and prevalence of falls prevention practice will be extracted. Qualitative data extraction will consist of themes or subthemes relevant to the review question, with verbatim quotes as support for the themes. Subsequently, the findings will each be assigned to a corresponding level of credibility (from unequivocal, credible, or not supported). These findings will be presented in tabular form, with narrative summaries as required.^(28)^

## Data transformation

The quantitative data will subsequently be converted into ‘qualitised data’. This will involve converting the quantitative data into textual descriptions or narrative summary where they will respond directly to the review questions.^(28,34)^ Through this process, data patterns with similar meanings will be identified during the data coding process, hence allowing similar codes to be grouped forming specific themes.^(35)^ Data will be coded against the phenomenon of interest in the inclusion criteria.

## Data Analysis and Synthesis

The review will use a convergent integrated approach according to the JBI methodology for mixed methods systemic reviews set out by JBI SUMARI.^(28)^ The qualitised and qualitative data will be systematically coded and categorised using thematic synthesis, hence developing descriptive themes and subthemes to create a Summary of Findings.^(36)^ Depending on the results, summaries will be created, using the WFG recommendations as a framework, for falls risk screening, providing advice about falls prevention and optometrists’ knowledge and awareness about the evidence for falls management. Analytical themes may be developed from the descriptive themes depending on the results.

## Ethics and dissemination

As no primary research will be conducted, ethical approval will not be necessary to conduct this review. The protocol is registered with PROSPERO (CRD42024539668) and the final manuscript will be published in a peer-reviewed journal.

## Strengths and limitations of the review

The included peer-reviewed studies will provide the best available evidence for understanding the practice patterns of optometrists in falls prevention management in community-settings. The review will only include optometrists as the participants, hence studies that investigate other ophthalmic professionals such as Opticians and Ophthalmologists will be excluded. Due to the inherent characteristics of a mixed-methods systematic review, the data will be integrated from diverse sources, which is recognised as a challenge in evaluating the quality of the evidence through conventional methods such as the GRADE or ConQual frameworks.^(28)^ This approach is discouraged in the JBI guidelines for mixed methods systematic reviews. However, including both quantitative and qualitative studies will provide a comprehensive synthesis of peer-reviewed literature. The results of the review will inform future research, and have a potential impact on policy-making, and optometric clinical practice concerning falls prevention management. The study selection criteria include studies written in English which may mean evidence presented in other languages is not included.

## Conclusion

Integrating falls prevention strategies within the optometry discipline has received limited attention within the existing literature. This mixed-methods systematic review will address the underexplored integration of falls prevention within optometry, aiming to synthesise the best available evidence about optometrists’ current practice patterns in implementing falls prevention management for older adults in community settings. The results of this review will provide valuable insights into the practice patterns of optometrist-related falls prevention management and how current falls guidelines. The findings will be useful in generating further education for optometrists and improving the public’s knowledge of falls prevention management.

## Contributions

SYL was responsible for the original idea for this review, designed the background and inclusion criteria, and wrote the first draft, with support from AMH. All authors reviewed and edited the draft and approved the final submission.

## Data Availability

All data produced in the present work are contained in the manuscript.

## Appendix 1

Example search Strategy

Ovid MEDLINE(R) ALL <1946 to May 14, 2024>

|  |  |
| --- | --- |
| 1 | (optometr* or ophthalmolog* or eyecare professional or eyecare practitioner or eye care profession* or eye care practitioner or eye health professional or eye health profession*).mp. |
| 2 | exp optometrists/ or optometry/ or ophthalmology/ |
| 3 | 1 or 2 |
| 4 | fall*.mp. |
| 5 | falling.mp. |
| 6 | exp Accidental Falls/ |
| 7 | 4 or 5 or 6 |
| 8 | exp aged/ or exp geriatrics/ or exp geriatric nursing/ or (centarian* or centenarian* or elder* or eldest or frail* or geriatri* or nonagenarian* or octagenarian* or octogenarian* or old age* or older adult* or older age* or older female* or older male* or older man or older men or older patient* or older people or older person* or older population or older subject* or older woman or older women or oldest old* or senior* or senium or septuagenarian* or supercentenarian* or very old*).mp. |
| 9 | exp Aged/ or "Aged, 80 and over"/ |
| 10 | 8 or 9 |
| 11 | 3 and 7 and 10 |
| 12 | limit 11 to yr="1980 - current" |

## Reference

1. Australian Institute of Health and Welfare. Falls. Canberra: AIHW; 2023 [cited 2024 07 May 2024]. Available from: https://www.aihw.gov.au/reports/injury/falls

2. Australian Institute of Health and Welfare. Falls in older Australians aged 65 and over 2019-20. Department of Health, editor.: Australian Institute of Health and Welfare; 2022. Available from: <u>https://</u>www.aihw.gov.au/getmedia/467fd7e8-bb5d-46b0-890d-c7235591804b/aihw-injcat-226-fact-sheet.pdf.aspx

3. Ganz DA, Latham NK. Prevention of Falls in Community-Dwelling Older Adults. The New England journal of medicine. 2020;382(8):734–743. doi:10.1056/NEJMcp1903252. Cited in: Pubmed; PMID cdi_proquest_journals_2358741697.

4. World Health Organization. Falls. World Health Organisation; 2021 26 April 2021. Available from: https://www.who.int/news-room/fact-sheets/detail/falls

5. Singh RR, Maurya P. Visual impairment and falls among older adults and elderly: evidence from longitudinal study of ageing in India. BMC Public Health. 2022;22(1):2324–2324. doi:10.1186/s12889-022-14697-2. Cited in: Pubmed; PMID cdi_webofscience_primary_000898481900003.

6. Killeen OJ, De Lott LB, Zhou Y, Hu M, Rein D, Reed N, et al. Population Prevalence of Vision Impairment in US Adults 71 Years and Older: The National Health and Aging Trends Study. JAMA Ophthalmology. 2023;141(2):197–204. doi:10.1001/jamaophthalmol.2022.5840.

7. Umfress AC, Brantley MA, Jr. Eye Care Disparities and Health-Related Consequences in Elderly Patients with Age-Related Eye Disease. Semin Ophthalmol. 2016;31(4):432–8. eng. Epub 2016/04/27. doi:10.3109/08820538.2016.1154171 [doi]. Cited in: Pubmed; PMID 27116323.

8. Klein R, Klein BE. The prevalence of age-related eye diseases and visual impairment in aging: current estimates. Invest Ophthalmol Vis Sci. 2013 Dec 13;54(14):ORSF5-ORSF13. eng. Epub 2013/12/18. doi:54/14/ORSF5 [pii]10.1167/iovs.13-12789 [doi]. Cited in: Pubmed; PMID 24335069.

9. Australian Institute of Health and Welfare. Injury in Australia: Falls. Australian Government, editor.: Australian Institute of Health and Welfare; 2023. Available from: https://www.aihw.gov.au/reports/injury/falls

10. Ramulu P, van Landingham S, Massof R, Chan E, Ferrucci L, Friedman D. Fear of Falling and Visual Field Loss from Glaucoma. Ophthalmology (Rochester, Minn). 2012;119(7):1352–1358. doi:10.1016/j.ophtha.2012.01.037. Cited in: Pubmed; PMID cdi_openaire_primary_doi_dedup_1b2c60a162588f4aa1661f23081609a6.

11. Keay L, Palagyi A, McCluskey P, Lamoureux E, Pesudovs K, Lo S, et al. Falls in Older people with Cataract, a longitudinal evalUation of impact and riSk: the FOCUS study protocol. Injury prevention. 2014;20(4):e7–e7. doi:10.1136/injuryprev-2013-041124. Cited in: Pubmed; PMID cdi_pubmed_primary_24431102.

12. Wood JM, Lacherez P, Black AA, Cole MH, Boon MY, Kerr GK. Risk of falls, injurious falls, and other injuries resulting from visual impairment among older adults with age-related macular degeneration. Investigative ophthalmology & visual science. 2011;52(8):5088–5092. doi:10.1167/iovs.10-6644. Cited in: Pubmed; PMID cdi_crossref_primary_10_1167_iovs_10_6644.

13. Johnson L, Buckley JG, Scally AJ, Elliott DB. Multifocal Spectacles Increase Variability in Toe Clearance and Risk of Tripping in the Elderly. Investigative ophthalmology & visual science. 2007;48(4):1466–1471. doi:10.1167/iovs.06-0586. Cited in: Pubmed; PMID cdi_pascalfrancis_primary_18652890.

14. Australian Government, Department of Health and Aged Care. Medicare Benefits Schedule - Item 10911. MBS Online Medicare Benefits Schedule: Australian Government, Department of Health and Aged Care; 2015 03 April 2024. Available from: https://www9.health.gov.au/mbs/fullDisplay.cfm?type=item&q=10911&qt=ItemID

15. Montero-Odasso M, Tan MP, Ryg J, Blain, H, Bourke R, Cameron ID, Camicioli R, et al. World guidelines for falls prevention and management for older adults: a global initiative. Age Ageing. 2022;51(9). doi:10.1093/ageing/afac205.

16. Ho KC, Elliott D, Charlesworth E, Gyawali R, Keay L. Feasibility of Implementing Recommendations to Reduce Fall Risk in Older People: A Delphi Study. Optometry and vision science. 2022;99(1):18–23. doi:10.1097/opx.0000000000001829. Cited in: Pubmed; PMID cdi_openaire_primary_doi_dedup_34f647c39b61c80dd5268603f376c4f8.

17. Miyawaki CE, Mauldin RL, Carman CR. The potential of optometrists’ referrals of older patients to community-based exercise programs: Findings from a mixed-methods study. Journal of aging and physical activity. 2020;28(2):194–207. doi:10.1123/japa.2018-0442. Cited in: Pubmed; PMID cdi_scopus_primary_2006870877.

18. Bako O. Eye exams can help prevent falls; Alberta optometrists and University of Alberta Injury Prevention Centre partner to raise awareness. The Westlock News. 2016. Cited in: Pubmed; PMID cdi_proquest_newspapers_1842497319.

19. Morgan DR. Optometrists can play significant role in fall prevention for older adults. Primary Care Optometry News. 2013;18(1):1. Cited in: Pubmed; PMID cdi_proquest_reports_1346848161.

20. Ming Y, Zecevic AA, Hunter SW, Miao W, Tirona RG. Medication Review in Preventing Older Adults’ Fall-Related Injury: a Systematic Review & Meta-Analysis. Canadian geriatrics journal CGJ. 2021;24(3):237–250. doi:10.5770/cgj.24.478. Cited in: Pubmed; PMID cdi_pubmedcentral_primary_oai_pubmedcentral_nih_gov_8390322.

21. Sherrington C, Whitney JC, Lord SR, Herbert RD, Cumming RG, Close JCT. Effective Exercise for the Prevention of Falls: A Systematic Review and Meta-Analysis. Journal of the American Geriatrics Society (JAGS). 2008;56(12):2234–2243. doi:10.1111/j.1532-5415.2008.02014.x. Cited in: Pubmed; PMID cdi_crossref_primary_10_1111_j_1532_5415_2008_02014_x.

22. Mak TCT, Wong TWL, Ng SSM. Visual-related training to improve balance and walking ability in older adults: A systematic review. Experimental gerontology. 2021;156:111612–111612. doi:10.1016/j.exger.2021.111612. Cited in: Pubmed; PMID cdi_webofscience_primary_000720659200005.

23. Close JCT, Hooper R, Glucksman E, Jackson SHD, Swift CG. Predictors of falls in a high risk population: results from the prevention of falls in the elderly trial (PROFET). Emergency medicine journal : EMJ. 2003;20(5):421–425. doi:10.1136/emj.20.5.421. Cited in: Pubmed; PMID cdi_webofscience_primary_000185103400012CitationCount.

24. Dillon L, Clemson L, Ramulu P, Sherrington C, Keay L. A systematic review and meta-analysis of exercise-based falls prevention strategies in adults aged 50+ years with visual impairment. Ophthalmic & physiological optics. 2018;38(4):456–467. doi:10.1111/opo.12562. Cited in: Pubmed; PMID cdi_crossref_primary_10_1111_opo_12562.

25. Elliott D, Black, A, Wood, J. Guidelines to help optometrists prevent falls in older patients. University of Bradford QUT, editor. Optometry Australia; 2019. [07 May 2024]. Available from: https://www.optometry.org.au/wp-content/uploads/Professional_support/Guidelines/Falls_Guidelines_v8.pdf

26. Ashleigh Chandra LK, Lisa Dillon, Jessie Huang, Iris Gordon. Vision impairment and eye-related interventions to reduce falls risk: a systematic review. PROSPERO; 2020 05 July 2020. PROSPERO [cited 2024 04 April 2024]. Available from: https://www.crd.york.ac.uk/prospero/display_record.php?RecordID=187617

27. Kingsley Ekemiri NE, Carl Abraham, Chioma Ekemiri, Osaze Okonedo, Virgina Victor, Robin Seemongal-Dass, Diane Van Staden. Global Burden of Fall and Associated Factor among Individuals with Low Vision: A systematic Review and Meta-Analysis. 2024 2024. doi:https://www.crd.york.ac.uk/prospero/display_record.php?RecordID=497563.

28. Lizarondo L, Stern C, Carrier J, Godfrey C, Rieger K, Salmond S, Apóstolo J, Kirkpatrick P, Loveday H. Chapter 8: Mixed Methods Systematic Reviews. JBI Manual for Evidence Synthesis. 2024. doi:10.46658/jbimes-20-09.

29. NLM. Optometrist MeSH Descriptor Data 2024. National Library of Medicine; 2018 05/02/2024. [cited 2024 05/02/2024]. Available from: https://meshb.nlm.nih.gov/record/ui?ui=D000072162

30. Aromatais E, Munn Z (Editor). JBI manual for Evidence Synthesis Adelaide, Australia: JBI; 2020 21 Feb 2024. Available from: 10.46658/JBIMES-20-01

31. Page MJ, McKenzie JE, Bossuyt PM, Boutron I, Hoffmann TC, Mulrow CD, Shamseer L, et al. The PRISMA 2020 statement: an updated guideline for reporting systematic reviews. BMJ. 2021 Mar 29;372:n71. eng. Epub 2021/03/31. doi:pagm061899 [pii]10.1136/bmj.n71 [doi]. Cited in: Pubmed; PMID 33782057.

32. Tufanaru C, Munn Z, Aromataris E, Campbell J, Hopp L. Chapter 4: Systematic reviews of effectiveness JBI Manual for Evidence Synthesis. 2020. doi:10.46658/JBIMES-24-03.

33. Lockwood CS, Porritt K, Munn Z, Rittenmeyer L, Salmond SW, Bjerrum MB, Loveday HP, Carrier J, Stannard D. Chapter 3: Systematic Reviews of Qualitative Evidence. JBI Manual for Evidence Synthesis. 2024 2024. doi:10.46658/JBIMES-24-02.

34. Frantzen KK, Fetters MD. Meta-integration for synthesizing data in a systematic mixed studies review: insights from research on autism spectrum disorder. Quality & Quantity. 2016;50(5):2251–2277. doi:10.1007/s11135-015-0261-6. Cited in: Pubmed; PMID Frantzen2016.

35. Heyvaert M, Maes B, Onghena P. Mixed methods research synthesis: definition, framework, and potential. Quality & Quantity. 2013;47(2):659–676. doi:10.1007/s11135-011-9538-6. Cited in: Pubmed; PMID Heyvaert2013.

36. Thomas J, Harden A. Methods for the thematic synthesis of qualitative research in systematic reviews. BMC Med Res Methodol. 2008 Jul 10;8:45. eng. Epub 2008/07/12. doi:1471-2288-8-45 [pii]10.1186/1471-2288-8-45 [doi]. Cited in: Pubmed; PMID 18616818.

